# Hypertension associated with the risk of extrahepatic cancers in MASLD population: a multicenter cross-sectional study in China

**DOI:** 10.1101/2025.07.28.25332237

**Authors:** Xinyue Zhao, Feng Xue, Shanshan Wang, Haiyun Ding, Dong Li, Huiying Rao, Fanpu Ji, Jidong Jia, Xiong Ma, Peng Hu, Xiaoguang Dou, Keshu Xu, Shuangqing Gao, Ming Yang, Lai Wei

## Abstract

**Background and aim:** Extrahepatic cancers have been recognized as a significant outcome of non-alcoholic fatty liver disease (NAFLD)--redefined as metabolic dysfunction-associated steatosis liver disease (MASLD) with five cardiometabolic risk factors including hypertension which is associated with several cancers’ tumorigenesis or anti-cancer treatment. We aimed to investigate the association between hypertension, liver fibrosis and extrahepatic cancers in MASLD population.

**Methods:** This multicenter cross-sectional study was based on MASLD population from hospital-based database across 11 centers in the nationwide of China, according to MASLD diagnostic criteria based on keywords and ICD-10 codes. Logistic regression was used to estimate odds ratio (OR) and 95% confidence interval (CI) between risk factors and extrahepatic cancers.

**Results:** Totally 103,652 MASLD individuals were identified, with 6,605 diagnosed extrahepatic cancers. The primary outcome showed that hypertension (OR 1.15, 95% CI: 1.05, 1.26), and its combination with hyperglycemia (OR 1.33, 95% CI: 1.19, 1.48) were significantly associated with extrahepatic cancers in MASLD population. Over 40-year-old, female gender, AST/ALT over ULN were risk factors. Metabolic-based treatments including ACEIs/ARBs (aOR 0.68, 95% CI:0.61,0.76), Fibrates (aOR 0.46, 95% CI:0.34,0.61), GLP-1 RAs (aOR 0.54, 95% CI:0.37,0.79), and Thiazolidinediones (aOR 0.45, 95% CI:0.26,0.79) were significantly protective factors. After adjusting confounding factors, FIB-4 index associated with extrahepatic cancers in all age groups. In hypertension subgroup, FIB-4 between 1.3-2.66 and over 3.48 were associated with extrahepatic cancers in aged 35 to 65, consistent with those over 65 with FIB-4 over 2.

**Conclusion:** Hypertension combined with liver fibrosis is associated with extrahepatic cancers in MASLD population.

## Introduction

The prevalence of non-alcoholic fatty liver disease (NAFLD) is currently estimated at around 38%^1^, with a predicted rise to 62.5% by 2040 in Asia^2^. Findings from population studies, community health screenings, and hospital-based databases have well established metabolic dysfunctional comorbidities and relevant outcomes of NAFLD population^3^. In 2020, metabolic dysfunction-associated fatty liver disease (MAFLD) was introduced, which firstly included metabolic syndrome, rather than exclusive diagnoses as NAFLD—excluding alcoholic intaking^4^. Subsequently, three major pan-national liver associations redefined metabolic dysfunction-associated steatosis liver disease (MASLD), which has been updated from non-alcoholic steatohepatitis (NASH) to metabolic dysfunction-associated steatohepatitis (MASH)^5^. This new nomenclature included five cardiometabolic risk factors: overweight or increased waist circumference, pre-diabetes and type 2 diabetes mellitus(T2DM), hypertension, hypertriglyceridemia and lower high-density lipoprotein cholesterol (HDL-C). The population affected by MASLD has been estimated to be consistent with that of NAFLD^6^.

Previous studies demonstrated that cardiovascular diseases, as the first extrahepatic fatal outcomes among chronic hepatic disease, were the leading cause-specific mortality among NAFLD population ^7,8^. However, extrahepatic cancers were identified another cause-specific mortality in NAFLD population, even regarded as first cause-specific mortality in some study^9^. The association between NAFLD and extrahepatic cancers has been identified over the past decades^10^. Meanwhile, obesity or overweight, along with T2DM are acknowledged risk factors of extrahepatic cancers in NAFLD populations^10^. Interestingly, in general cancers population, hypertension has been identified as comorbidity during anti-cancer treatments or risk factors of cancers^11^.

Though emerging evidence has shown strong association between MASLD and an increased risk of extrahepatic cancers ^10^, regarding to five cardiometabolic risk factors, especially hypertension, their association with extrahepatic cancers in MASLD population was unclear. Therefore, we hypothesize that hypertension is associated with the risk of extrahepatic cancers in the MASLD population. The combinations of those, including widely acknowledged diabetes or dyslipidemia, metabolic-based treatments, and the severity of liver fibrosis, will be illustrated in this study as well.

## Methods

### Population and study design

This multicenter cross-sectional retrospective research based on a multicenter hospital-based database, from 11 tertiary hospitals across 6 cities in China (Beijing, Shanghai, Xi’an, Chongqing, Shenyang, and Wuhan), which was compiled between 1/ 1/2020 and 12/ 31/2022.

### Definition

#### Definition of MASLD

According to the criteria for the diagnosis of MASLD proposed in 2023^12^. MASLD was diagnosed (≥18-year-old) as evidence of hepatic steatosis, with one of the following five cardiometabolic criteria, (I) overweight/obesity [body mass index (BMI) ≥23.00 kg/m^2^ for Asians] OR waist circumference≥94cm(male) 80cm(female), (II) Fasting serum glucose ≥5.6 mmol/L [100 mg/dl] OR 2-hour post-load glucose levels ≥7.8 mmol/L [≥140 mg/dl] OR HbA1c ≥5.7% [39 mmol/L] OR type 2 diabetes OR treatment for type 2 diabetes mellitus, (III) Blood pressure ≥130/85 mmHg OR specific antihypertensive drug treatment, (IV) Plasma triglycerides ≥1.70 mmol/L [150 mg/dl] OR lipid lowering treatment, (V) Plasma High density lipoprotein cholesterol ≤1.0 mmol/L [40 mg/dl] (male) and ≤1.3 mmol/L [50 mg/dl] (female) OR lipid lowering treatment.

#### Definition of Hyperlipidemia

When ≥1 of the following fasting venous plasma test indicators is met: total cholesterol (TC) ≥5.2 mmol/L; Low-density lipoprotein cholesterol (LDL-C) ≥3.4 mmol/L; Triglyceride (TG) ≥1.7 mmol/L; High-density lipoprotein cholesterol (HDL-C) < 1.0 mmol/L for males and <1.3 mmol/L for females, named lower HDL-C in followings^13^.

### Ascertainment of MASLD population and extrahepatic cancers

The accuracy of diagnosis has been evaluated by experienced clinicians through random extractions according to ICD-10 codes. Firstly, those diagnosed “hepatic steatosis” were extracted from the database using ICD-10 codes with keywords (in Chinese), excluding others etiological hepatic steatosis. The ICD-10 codes and keywords (in Chinese) to identify the MASLD population were illustrated in Supplementary Table 1. Screening and appraisal to identify and extract those who met the MASLD diagnostic criteria were done. According to real-world diagnostic routines, the MASLD cardiometabolic criteria (IV) included the diagnosis of ‘Hyperlipidemia’. The absence of body mass index (BMI) and waistline circle (WC) (I) in our database meant that these two criteria were not utilized. Based on these criteria, patients diagnosed with carcinomas were identified through diagnostic records. Finally, a total of 103,652 MASLD patients were included in this study (Figure 1).

**Figure 1:**
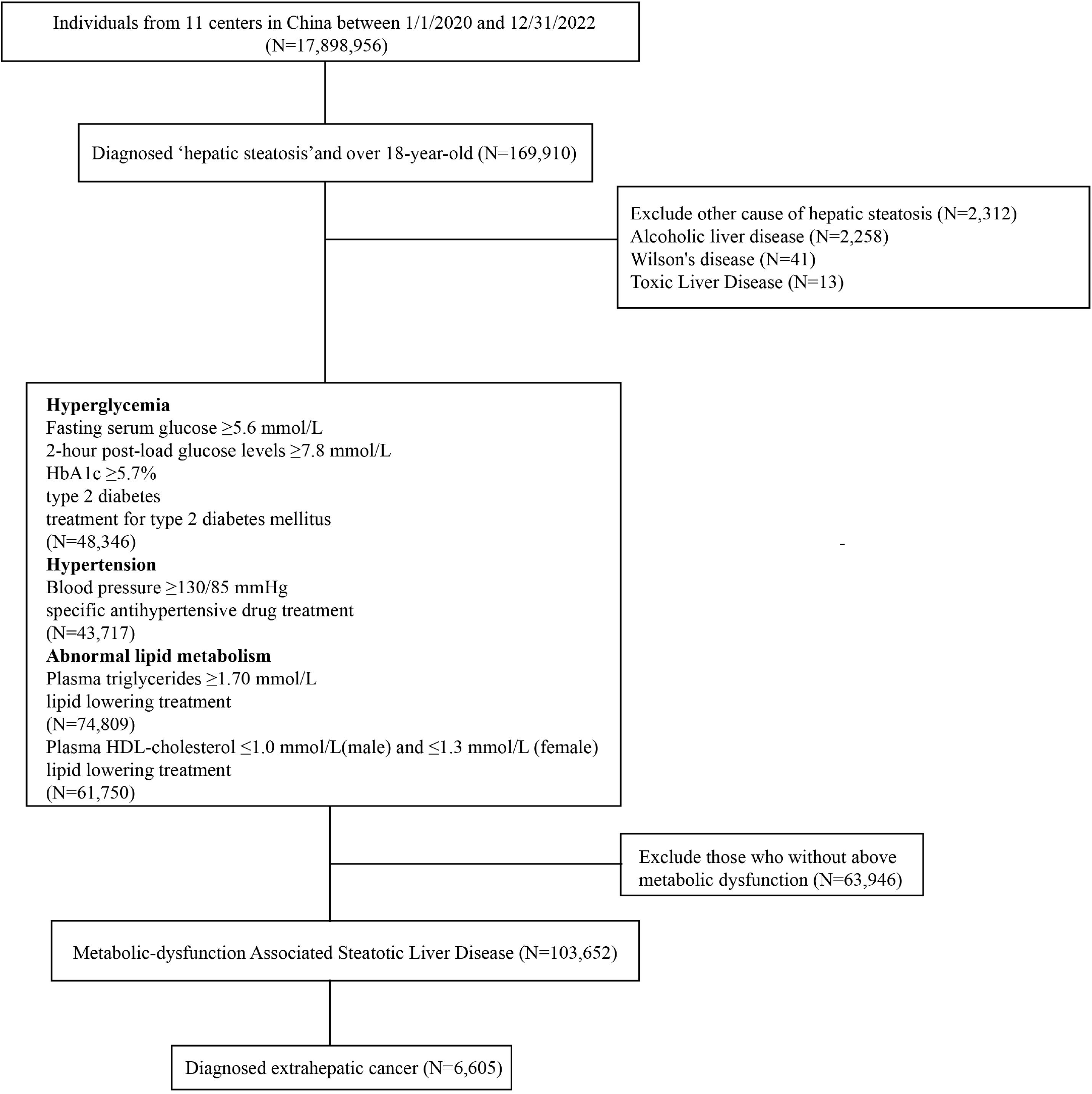
The flow chart of the screening of extrahepatic cancer in MASLD population.

### Subgroups

In logistic regressive analysis, we classified metabolic dysfunctions as following: hypertension, including diagnosed hypertension and use anti-hypertension agents; abnormal lipid metabolism, including plasma triglycerides over 1.70 mmol/L and diagnosed hyperlipidemia, lower HDL-C and using lipid lowering treatment; hyperglycemia, pre-diabetes: fasting blood-glucose 5.6-6.9mmol/L, HbA1c 5.7-6.4%, 2-hour post-meal blood glucose 7.8-11.0mmol/L, T2DM and treatment for T2DM. In metabolic dysfunctional subgroup analysis including, hypertension group (diagnosed hypertension and use anti-hypertension agents), abnormal lipid metabolism group (plasma triglycerides over 1.70 mmol/L, diagnosed hyperlipidemia, lower HDL-C and lipid lowering treatment), and T2DM group (diagnosed T2DM and treatment for T2DM). Ascertainment of pharmacological treatments of the metabolic dysfunction and comorbidities were shown in supplementary materials.

### Fibrosis assessment

The Fibrosis-4 index (FIB-4) ^14,15^ was calculated using the established equation that incorporates age, AST level, ALT level, and platelet count. In individuals aged 35 to 65 years^16^, FIB-4 of less than 1.3 (F0–F1) was considered low risk for advanced fibrosis, while a score ranging from 1.3 to 2.67 (F2) indicated intermediate risk and required further evaluation through liver stiffness measurement via elastography, liver function tests, or other methods. The score exceeding 2.67 and 3.48^17^ was classified as high risk for advanced fibrosis (F3–F4) and cirrhosis, which linked to an increased risk of adverse liver outcomes. For those over 65 years old, the FIB-4 cutoff was raised to 2^16^.

### Statistical analysis

SAS Version 9.4 (SAS Institute Inc., Cary, NC, USA) was used for statistical analysis, with p<0.05 as significant difference. Continuous variables were expressed as mean ± standard deviation (SD); values were presented as the median and interquartile range (Q1, Q3) and analyzed using the Wilcoxon Test or Kruskal-Wallis Test for skewed distributions. Univariate and multivariate logistic regression analyses were conducted to evaluate the risk factors for extrahepatic cancers, with the odds ratio (OR), adjusted odds ratio (aOR) and 95% confidence interval (95% CI) calculated. Covariate were identified according to clinical importance and statistical significance. Considering clinical impact and statistical results in univariate logistic analysis, multivariate logistic regression analysis was done. Forest graphs were made by R 4.4.2.

## Result

### Demographic, clinical characters, and division of extrahepatic cancers in MASLD population

A total of 6,605 MASLD individuals with extrahepatic cancers were identified, with 3,243 (49.10%) females. The mean age was 58.61 ± 12.49. Dyslipidemia was the most prevalent at 4,275(64.72%). Together with the combination of cardiometabolic risk factors and other basic clinical characteristics including comorbidities and metabolic-based treatments were presented in Supplementary Table 3. As the absence of weight and waistline in our database, those two was not included in descriptive statistics and logistic regressive analysis. Overall, lung cancer, thyroid cancer, and breast cancer were identified as the top three extrahepatic malignant carcinomas. Gender difference of site-specific extrahepatic cancers in our MASLD population are detailed. There was no difference in incidence between genders (Figure2, Supplementary Table 4).

### Hypertension associated with extrahepatic cancers in MASLD population

The univariate logistic analysis indicated that ageing, female gender, and cardiometabolic risk factors were significantly linked to extrahepatic cancers in MASLD population (Supplementary Table 5). Considering the interaction of risk factors, multivariate logistic regression analysis was performed.

Hypertension was significantly associated with the risk of extrahepatic cancers in the MASLD population (OR 1.15, 95% CI: 1.05, 1.26). As for age, compared to under 30-year-old, individuals over 40-year-old associated over threefold possibility of developing extrahepatic cancers(P<0.05). Additionally, female gender (OR 1.41, 95% CI: 1.34, 1.48), alanine aminotransferase (ALT) (OR 1.24, 95% CI: 1.13, 1.36), aspartate aminotransferase (AST) (OR 1.18, 95% CI: 1.09, 1.28) over 40 U/L were associated with a higher risk of extrahepatic cancers (in Figure 3). Furthermore, the combination of hypertension and hyperglycemia (OR 1.33, 95%CI:1.19,1.48) was significantly linked to the risk of extrahepatic cancers in the MASLD population (Figure 4). Specifically, as shown in Supplementary Figure 1, diagnosed hypertension (OR 1.14, 95% CI: 1.08, 1.21) and T2DM (OR 1.15, 95% CI: 1.08, 1.22) were risk factors for extrahepatic cancers. In specific dyslipidemia, none of them was identified (Supplementary Figure 2).

**Figure 2:**
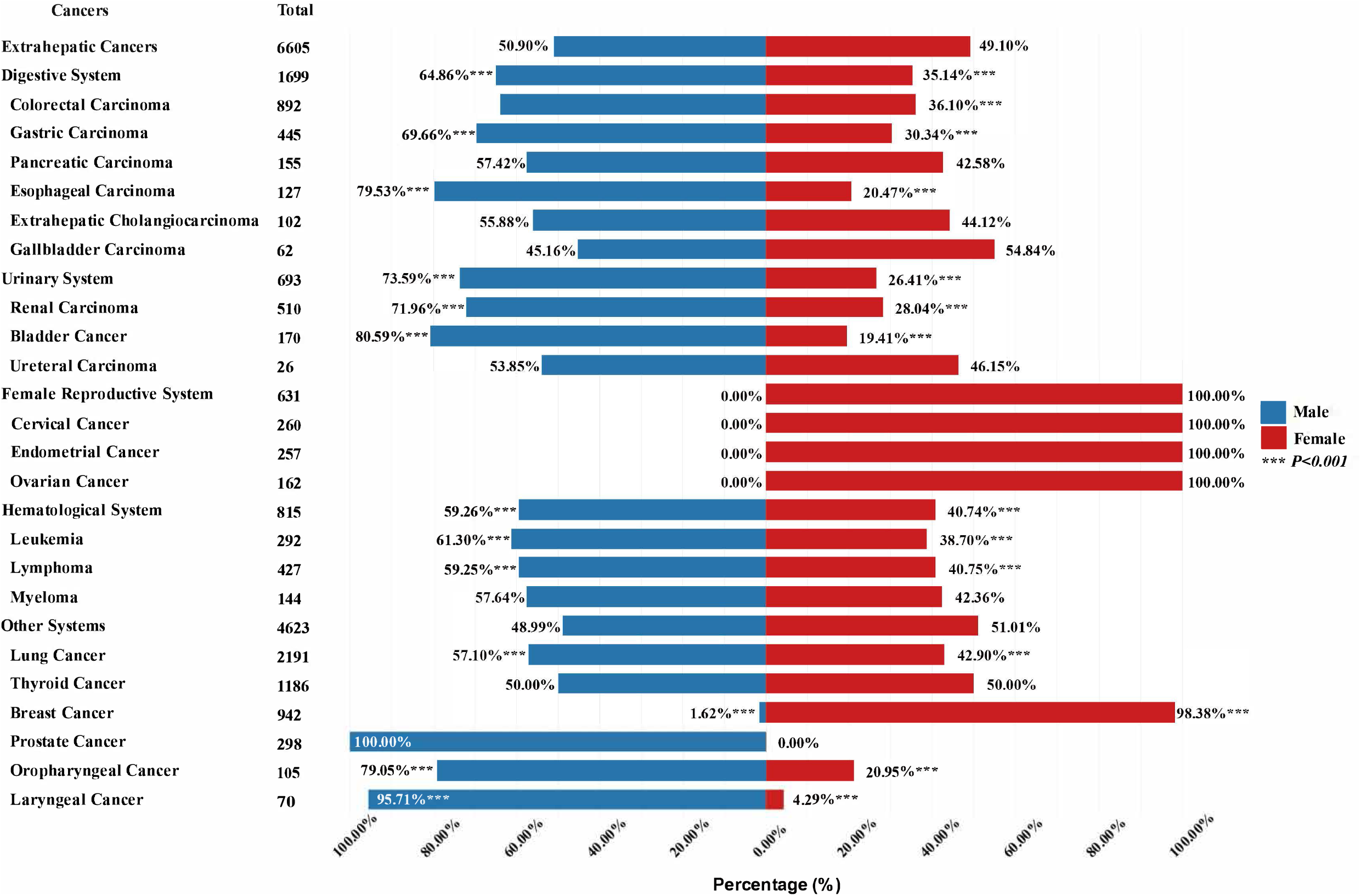
Gender difference of extrahepatic cancers in MASLD population

**Figure 3:**
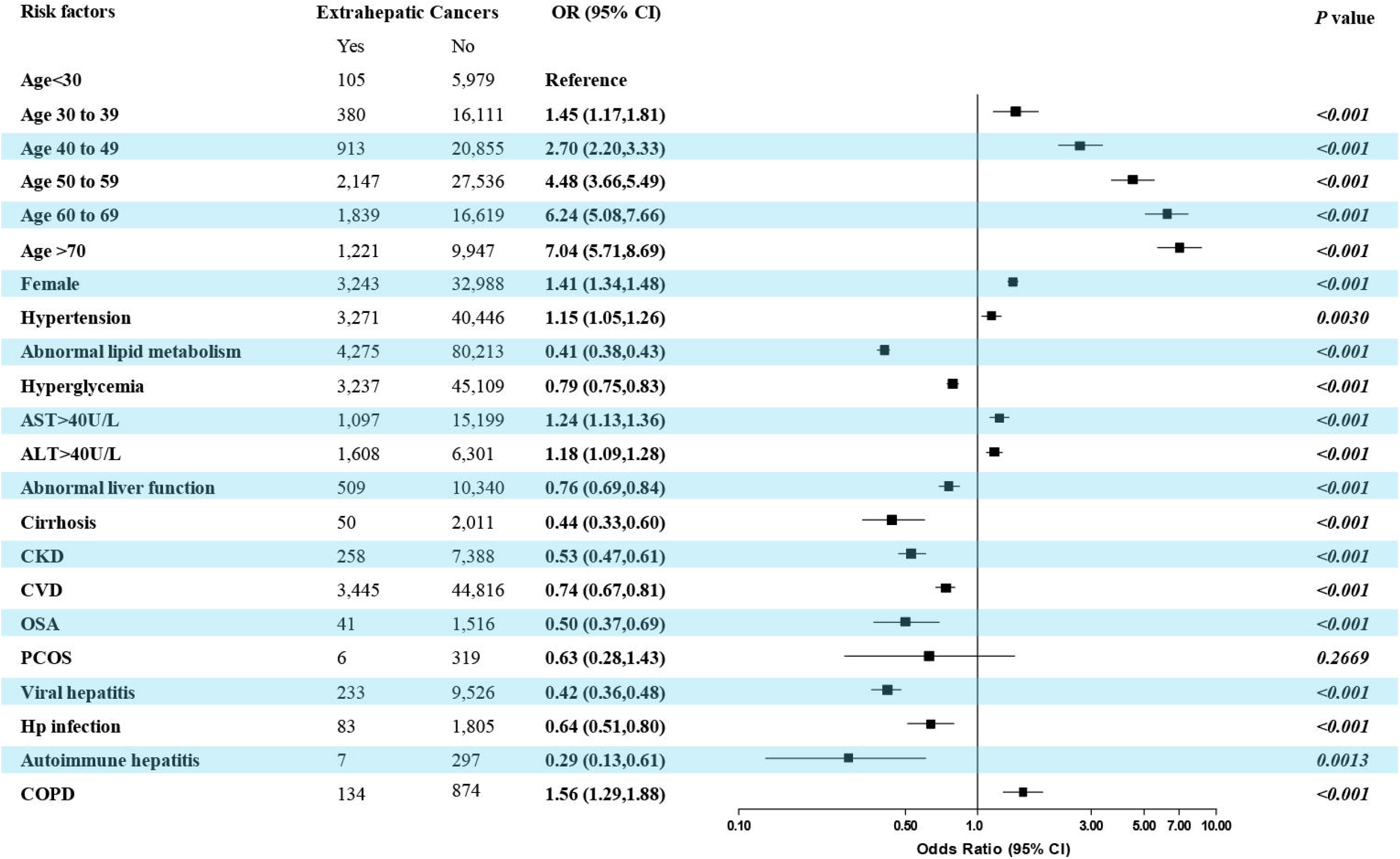
The association of metabolic dysfunctions, elevated liver enzyme, comorbidities and extrahepatic cancers in the MASLD population. ALT, Alanine Aminotransferase, AST, Aspartate Aminotransferase, CKD, Chronic kidney disease, OSA, Obstructive Sleep Apnea, PCOS, Polycystic Ovarian Syndrome, COPD, Chronic obstructive pulmonary disease.

**Figure 4:**
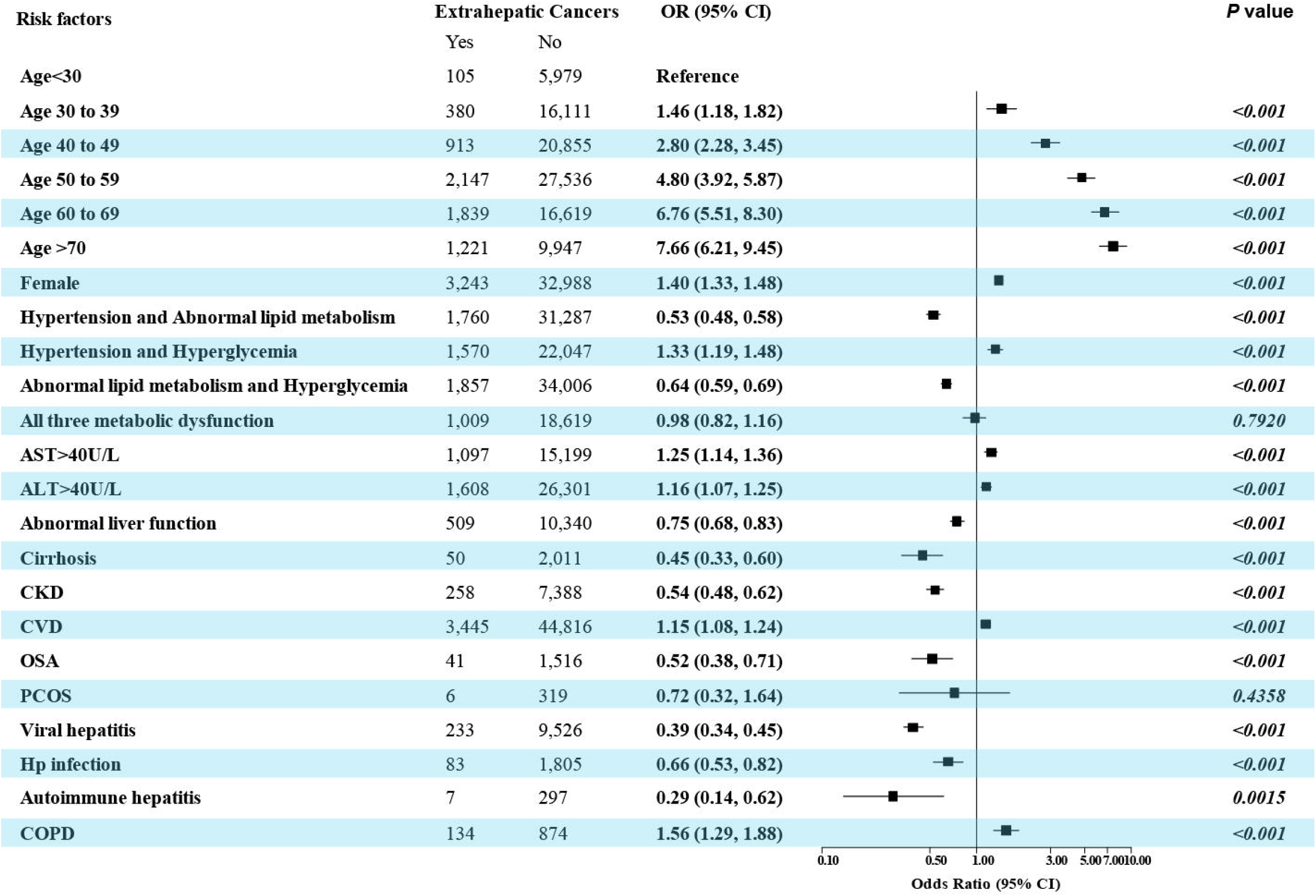
The association of the combination of cardiometabolic dysfunction factors, elevated liver enzyme, comorbidities and extrahepatic cancers in the MASLD population. ALT, Alanine Aminotransferase, AST, Aspartate Aminotransferase, CKD, Chronic kidney disease, OSA, Obstructive Sleep Apnea, PCOS, Polycystic Ovarian Syndrome, COPD, Chronic obstructive pulmonary disease.

### Pharmacological treatments of hypertension showed protective impact on extrahepatic cancers in MASLD population

Pharmacological treatments were protective factors as shown in Supplementary Figure 1 and 2. To confirm which category of agents protects MASLD population from extrahepatic cancers, subgroup analysis was conducted, among ‘hypertension’, ‘abnormal lipid metabolism’, and ‘T2DM’.

After adjusting confounding (supplementary tables 6-8), all forms of agents acted as protective factors in univariate analyses. In multivariate analysis, angiotensin-converting enzyme inhibitors/angiotensin II receptors (ACEIs or ARBs) (aOR 0.68, 95%CI:0.61,0.76) and Beta-Blockers (aOR 0.78, 95%CI:0.67,0.90) were protective factors. As for lipid-modifying agents, Fibrates demonstrated the highest protective qualification (aOR 0.46, 95%CI:0.34,0.61), Statins (aOR 0.62, 95%CI: 0.56,0.70) and Cholesterol Absorption Inhibitors (aOR 0.65, 95%CI: 0.47,0.91) exhibited protective capacity in our study. In T2DM, glucagon-like peptide-1 receptor agonists (GLP-1 RAs) (aOR 0.54, 95%CI:0.37,0.79) and Thiazolidinediones (aOR 0.45, 95%CI:0.26,0.79), showed on top two protective qualifications with lower usage rates. Insulin, sodium-glucose cotransporter-2 inhibitors (SGLT-2i), and dipeptidyl peptidase-4 inhibitors (DPP-4i) exhibited the same pattern. Metformin, though mostly used, was only associated with 13% lower protective possibilities.

### Fibrosis 4 index--associated with extrahepatic cancers in MASLD population

After adjusting confounding (supplementary tables 6-8), multivariate logistic regression analysis was conducted (presented in Table 2). In MASLD population, individuals aged 35 to 65, compared with FIB-4<1.3, FIB-4 over 1.3 showed a higher potential for developing extrahepatic cancers. The risky likelihood increased in FIB-4 1.3-2.66 (aOR 1.47, 95% CI: 1.25, 1.46), over 2.67 (aOR 1.47, 95% CI: 1.21, 1.78), and 3.48 (aOR 1.46, 95% CI: 1.27, 1.68), respectively. However, when compared with FIB-4 1.3-2.66, only FIB-4<1.3 acted as a protective factor with significant difference. In all subgroups, the MASLD population with FIB-4 over 1.3 was identified higher potential for developing extrahepatic cancers. Specifically, in hypertension subgroup, individuals aged 35-65 with FIB-4 1.3-2.66 and over 3.48, outnumbered 46% and 57% likelihood developing extrahepatic cancers compared with FIB-4<1.3, as well as T2DM subgroup (50% and 95%). As for abnormal lipid metabolism subgroup, individuals aged 35-65 with FIB-4 over 2.67 and 3.48 exhibited over two-fold. All of over 65- year-old with FIB-4 over 2 showed a higher potential for developing extrahepatic cancers.

## Discussion

Hypertension and its combination with hyperglycemia have been recognized as risk factor for developing extrahepatic malignancies. Generally, hypertension posed the greatest burden in 2021, particularly in India, China, and the United States^18^. In MASLD population, besides the understanding of the association between hypertension and the cardiac outcomes, others outcomes including malignancies or liver decompensation were insufficient. In the limited researches on the association of hypertension and cancers, though with arguments, hypertension is the risk factor of breast cancer in postmenopausal women^19^, thyroid, esophageal, colorectal, liver, and renal cell cancer ^11,20,21^. A system review and meta-analysis on risk factors of hepatocellular carcinoma (HCC) in NAFLD population unveiled that hypertension was risk factor of HCC with odds ratio at 1.75^22^. According to our results, though with only 15% higher risk possibilities, our study specifically identified the combination of hypertension and hyperglycemia, the odds ratio rise to 1.33 with significant difference. Based on the Kailuan cohort diagnosed with MAFLD^23^, metabolic dysfunction increased the risk of extrahepatic cancers, while the presence of two or three factors further heightened this risk. It might stem from the incidence of extrahepatic cancers in our MASLD population, which indicated that the combination of steatotic liver and hypertension or others metabolic-dysfunctional factors might associate with or even increase the risk of malignancies. Thus, diligent blood pressure monitoring should consistently be prioritized to address the potential risk of malignancies. In the context of hyperglycemia or dyslipidemia, numerous studies have highlighted that abnormal glucose and lipids associated with the increased risk of the progression of MASLD, fibrosis^24^, cardiac outcomes, and liver metastatic malignancies ^25^.

The impact of metabolic-base treatments has been illustrated in our study. This strongly suggests that etiological interventions may effectively prevent the emergence of malignancies in the MASLD population, which previous studies have proved that effectively slow the progression of MASLD and potentially prevent cancers^12,17^. Unfortunately, the global hypertension profile published by the World Health Organization (WHO) revealed a concerning trend in hypertension control rates—treatment rates were 39%, and only 16% met treatment compliance^26^. There is a gap between diagnosis and regular treatment. According to the results of previous clinical trial and observational studies, GLP-1 RAs have emerged as populate treatment in the fields of diabetes and obesity, with recent clinical trials unveiling significant benefits in the management and treatment of MASLD^27^, especially in those obese or hyperglycemia MASLD population. As for the management of dyslipidemia, statin was identified protective effect on overall mortality and cancer related mortality but are underutilized in NAFLD population^28^. According to our results, ACEIs/ARBs, Fibrates, GLP-1 RAs and Thiazolidinediones showed outstanding protective impact, which were recommended to alleviate metabolic dysfunction and regarded as protective factors on the emergence of malignancies in the MASLD population.

In the aspect of liver fibrosis, FIB-4 greater than or equal to 1.3 or 2 significantly associated with the risk of extrahepatic cancers in our study, which is consistent with a recent study in adults with MASLD that FIB-4 over 2.67 was associated with a 16% greater risk^29^. To our limited knowledge on liver fibrosis estimated via FIB-4 index in MASLD with hypertension population, the association of liver fibrosis and extrahepatic malignancies in MASLD population with hypertension was firstly demonstrated. Dyslipidemia has been illustrated that associated with the progression of MASLD, fibrosis, cardiac outcomes, or predict liver metastasis in MASLD population^25^. In our study, not only in T2DM subgroup which was consistent with previous study^30^, but dyslipidemia subgroup, all FIB-4 cut-offs showed as risk factors of extrahepatic cancers. Therefore, assessing liver fibrosis through either invasive or non-invasive methods is crucial for health screening and hospital care.

In the perspective of age, over 40-year-old experienced extreme surge of odds ratio, which meet the issue that MASLD significantly increased the risk of early-onset cancers ^31^.In our study, the female gender was risk factor. However, there’s no gender difference of the amounts of extrahepatic cancers which might stem from gender composition in various cancers in our database. Limited female MASLD population study revealed that women have higher risk of advanced liver fibrosis and cancers than men^32,33^. We suggest that those MASLD population should take participant in cancer screening in all age range, especially for those over 40-year-old women.

We acknowledge that there are several limitations in this study. This cross-sectional study excluded causality and follow-up with only associations demonstrated. The absence of diagnoses for obesity and overweight and data on alcohol consumption makes those factors could not be analyzed. However, apart from those, the participants in our MASLD population could meet the objectives of our research. Interestingly, it is MASLD, rather than obesity or overweight, that is independently associated with the incidence of malignancies^34^. Furthermore, the obesity paradox in MASLD patients with extrahepatic cancers has been unveiled^35^. Subsequent investigation on obesity in indicating cancer is still required.

The strengths of our study lie in the significant demonstration of each cardiometabolic risk factor, except for obesity, alongside liver fibrosis measured by FIB-4. Hypertension, and its combination with hyperglycemia were identified as risk factors associated with extrahepatic cancers. There have been few reports confirming specific single or combination of cardiometabolic risk factors potentially associated with extrahepatic cancers. Furthermore, subgroup analyses revealed protective metabolic-based treatment and the possibility of FIB-4 being used in cancer screening in MASLD population.

## Conclusion

A total of 6,605 individuals with extrahepatic cancers were identified within the MASLD population. Hypertension, the combination of hypertension and hyperglycemia, elevated liver enzymes, were significantly associated with extrahepatic cancers. Fortunately, metabolic-based treatments were significantly linked to a protective role in the MASLD population. ACEIs/ARBs, Fibrates, GLP-1 RAs and Thiazolidinediones were recommended. Managing metabolic dysfunction was not only revers the progression of MASLD, liver fibrosis, also prevent MASLD population from the risk of extrahepatic cancers. FIB-4 of over 1.3 for individuals aged 35 to 65, and over 2 for those aged over 65, was risk factors associated with extrahepatic cancers in each cardiometabolic risk factors subgroup. Specifically, hypertension combined with liver fibrosis is associated with extrahepatic cancers in MASLD population.

## Data Availability

All data produced in the present study are available upon reasonable request to the authors

## Declarations

## Acknowledgments

The authors thank Shuangqing Gao, Haiyun Ding, Shanshan Wang, Feng Xue, Huiying Rao, Fanpu Ji, Jidong Jia, Xiong Ma, Peng Hu, Xiaoguang Dou, Keshu Xu for data access and statistical analysis. Critically reviewing conducted by Lai Wei, Ming Yang, Dong Li, Shuangqing Gao, Huiying Rao, Fanpu Ji, Jidong Jia, Xiong Ma, Peng Hu, Xiaoguang Dou and Keshu Xu for scientific content.

## Author Contributions

Study concept and design: Xinyue Zhao and Lai Wei. Data acquisition: Haiyun Ding, Shanshan Wang, Shuangqing Gao, Feng Xue, Fanpu Ji, Jidong Jia, Huiying Rao, Xiong Ma, Peng Hu, Xiaoguang Dou, and Keshu Xu. Data analysis: Haiyun Ding, Shanshan Wang, Xinyue Zhao. Manuscript draft: Xinyue Zhao, Lai Wei. Data interpretation, critical review and revision of manuscript: Lai Wei, Ming Yang, Shuangqing Gao, Dong Li, Fanpu Ji, Jidong Jia, Huiying Rao, Xiong Ma, Peng Hu, Xiaoguang Dou, and Keshu Xu. Overall study supervision: Lai Wei. All authors participated in the preparation of the manuscript and have seen and approved the final version.

## Ethical Statement

This study was reviewed and approved by Beijing Tsinghua Changgung Hospital Ethics Committee, which waived the need for informed consent since the study only used deidentified databases, ID for ethics approval: 25469-0-01. We will adhere to all recommendations and requirements set forth by the board.

## Data Sharing Statement

The data were used under license for the current study and are therefore not publicly available.

