## Supplemental files for "Hypertension associated with the risk of extrahepatic cancers in MASLD population: a multicenter cross-sectional study in China"

**Supplementary materials content**

Supplementary Table 1. ICD-10 codes and keywords for cardiometabolic factors, comorbidities and cancers

| Disease | Code | Keywords |
|---|---|---|
| Hepatic Steatosis | K76 | Fatty Liver, Fatty Liver Inflammation, Nonalcoholic Steatohepatitis (NASH), Fatty Liver Disease, Fatty Liver Cirrhosis,Chronic Hepatitis, Unclassified Elsewhere, |
| Excluded Hepatic Steatosis | K70-K75, K77 | Alcoholic Steatohepatitis, Alcoholic Fatty Liver, Alcoholic Steatohepatitis , Alcoholic Fatty Liver Disease, Alcoholic Fatty Liver Cirrhosis; Toxic liver disease; Wilson's disease |
| Cardiometabolic Disease | | |
| Type 2 Diabetes Mellitus | E11 | Diabetes Mellitus, Glycosuria, Type 2 Diabetes, Type 2 Diabetes Mellitus, T2D, DM, T2DM |
| Hypertension | I10-I15 | Hypertension |
| Diagnosis Of Hyperlipidemia | E78 | High Cholesterol, Hyperlipidemia, Elevated Lipids, Cholesterolemia, Elevated Blood Lipids, Hypercholesterolemia, Cholesterol Syndrome, Mildly Elevated Blood Lipids, Hyperlipoproteinemia, High Cholesterol, Elevated Blood Lipids, Hyperlipemia, Hypertriglyceridemia, Dyslipidemia, Hyperlipidemia, High Triglycerides, Abnormal Blood Lipids, Abnormal Lipid Metabolism, Triglyceridemia, Lipid Metabolism Disorder, Lipoprotein Metabolism Disorder, Hyperlipidemia, Lipid Metabolism Disorder, Lipoprotein Deficiency, Lipid Metabolism Disorder |
| Comorbidities | | |
| Cardiovascular Disease | I20-I25, I30-I52 | All mentioned cardiovascular disease |
| Abnormal Liver Function | R90-R94 | Abnormal Results of Liver Function Tests |
| Viral Hepatitis | B15-B19 | Viral Hepatitis, Acute Hepatitis A, Acute Hepatitis B, Other Acute Viral Hepatitis, Chronic Viral Hepatitis, Unspecified Viral Hepatitis, Hepatitis A, Hepatitis B |

| | | |
|---|---|---|
| Chronic Kidney Disease | N03-N05, N11, N18, N19, I12, I13, D59.3, P96.0, R39.2, O10.2, O10.3 | Chronic Kidney Disease, Chronic Pyelonephritis, Hypertensive Nephropathy, Chronic Renal Insufficiency, Nephrotic Syndrome, Diabetic Nephropathy, Chronic Renal Failure, Chronic Kidney Wind (Note: This term may not have a direct medical equivalent in English), Diabetic Kidney Disease, Chronic Renal Failure, Nephritis Syndrome, Chronic Kidney Disease due to Diabetes, Chronic Kidney Failure, Renal Failure, Diabetic Kidney, Chronic Renal Function Decline, Renal Failure, Dialysis, Chronic Nephritis, Renal Failure, Hemodialysis, Chronic Glomerulonephritis, Kidney Disease Stage III, Hemodialysis, Chronic Kidney Damage, Chronic Kidney Disease, Hypertensive Kidney Disease |
| Osteoporosis | M80-M85 | Osteoporosis without Pathological Fractures, Osteoporosis with Pathological Fractures, Osteoporosis Due to Diseases Classified Elsewhere |
| Hypothyroidism | E00-E07 | Subclinical Iodine Deficiency Hypothyroidism with Mild Clinical Symptoms, Drug-Induced and Other Exogenous Hypothyroidism, Other Hypothyroidism |
| Cirrhosis | K70-K77 | Liver Cirrhosis, Liver Fibrosis and Hardening |
| Obstructive Sleep Apnea Syndrome | G47.3 | Sleep Apnea, Obstructive Sleep Apnea, Obstructive Sleep Apnea Syndrome |
| Polycystic Ovarian Syndrome | E28.2 | Polycystic Ovarian Syndrome |
| Chronic Obstructive Pulmonary Disease | J44 | Chronic obstructive pulmonary disease |
| All Cancer | | |
| Hepatic Carcinoma | | |
| Hepatocellular Carcinoma | C22.0 | Liver cancer, Hepatocellular carcinoma, Small cell liver carcinoma, HCC, Malignant liver tumor, Liver CA, Cholangiocarcinoma, Malignant cholangiocarcinoma, Intrahepatic cholangiocarcinoma, Malignant intrahepatic cholangiocarcinoma, Extrahepatic cholangiocarcinoma, Malignant hepatic cell tumor, Primary malignant liver tumor, Malignant tumor of the liver, Moderately differentiated mass-forming cholangiocarcinoma, Poorly differentiated hepatocellular |

| | | |
|---|---|---|
| Intrahepatic Cholangiocarcinoma | C22.1 | Intrahepatic bile duct cancer, Malignant intrahepatic bile duct tumor, Intrahepatic bile duct CA, Intrahepatic bile duct carcinoma, Malignant intrahepatic bile duct carcinoma, Intrahepatic bile duct cell CA, Intrahepatic bile duct adenocarcinoma, Intrahepatic bile duct adenocarcinoma CA, Intrahepatic bile duct small cell carcinoma, Malignant intrahepatic bile duct small cell tumor. |
| Extrahepatic Carcinoma<br>Digestive System Neoplasm | | |
| Colorectal Carcinoma | C18-C20 | Colorectal cancer, Colon cancer, Rectal cancer, Malignant colon tumor, Moderately differentiated adenocarcinoma of the colon, Malignant colon tumor, Cecal cancer, Malignant tumor of the cecum, Tubular adenocarcinoma of the cecum, Poorly differentiated ulcerative adenocarcinoma of the cecum, Mucinous adenocarcinoma of the cecum, Adenocarcinoma of the cecum, Moderately differentiated adenocarcinoma of the cecum, Descending colon CA, Descending colon cancer, Malignant tumor of the descending colon, Tubular adenocarcinoma of the descending colon, Moderately differentiated adenocarcinoma of the descending colon, Colon CA, Colon adenocarcinoma, Colonmalignant tumor, Appendiceal cancer, Malignant appendiceal tumor, Appendiceal adenocarcinoma, Cecal cancer, Malignant cecal tumor, Ascending colon adenocarcinoma, Ascending colon CA, Ascending colon cancer, Malignant tumor of the ascending colon, Cecal cancer, Malignant tumor of the cecum, Descending colon cancer, Malignant tumor of the descending colon, Moderately differentiated tubular adenocarcinoma of the descending colon, Colon CA, Hepatic flexure cancer, Malignant rectal tumor, Rectal CA, Rectal adenocarcinoma, Mucinous adenocarcinoma of the rectum |

| | | |
|---|---|---|
| Gastric Carcinoma | C16 | Gastric cancer, Fundic cancer, GC, Malignant gastric tumor, Gastric CA, Cardia cancer, Malignant tumor of the cardia, Cardia CA, Cardiac adenocarcinoma, Antrum cancer, Malignant tumor of the antrum, Poorly differentiated adenocarcinoma of the antrum, Body cancer of the stomach, Malignant tumor of the gastric body, Malignant fundic tumor, Gastric adenocarcinoma, Antral adenocarcinoma, Poorly differentiated adenocarcinoma of the stomach, Gastric tumor, Malignant, Cardia tumor |
| Pancreatic Carcinoma | C25 | Pancreatic cancer, Pancreatic carcinoma, Malignant tumor of the pancreatic neck, Malignant pancreatic tumor, Malignant pancreatic neoplasm, Body of the pancreas cancer, Malignant tumor of the pancreatic body, Head of the pancreas CA, Head of the pancreas cancer, Malignant tumor of the pancreatic head, Malignant tumor of the pancreatic tail, Pancreatic CA. |
| Esophageal Carcinoma | C15 | Esophageal cancer, Esophageal CA, Esophagus CA, Malignant esophageal tumor, Malignant tumor of the esophagus, Esophageal squamous cell carcinoma, Esophageal adenocarcinoma, Squamous cell carcinoma of the esophagus, Adenocarcinoma of the esophagus, Undifferentiated carcinoma of the esophagus, Small cell carcinoma of the esophagus, Undifferentiated esophageal carcinoma, Small cell esophageal carcinoma. |
| Extrahepatic Cholangiocarcinoma | C24.0 | Extrahepatic bile duct cancer, Malignant extrahepatic bile duct tumor, Extrahepatic bile duct CA, Extrahepatic bile duct carcinoma, Extrahepatic bile duct adenocarcinoma, Hilar bile duct CA, Hilar bile duct CA, Hilar bile duct cancer, Malignant hilar bile duct tumor, Malignant common bile duct tumor, Malignant hepatic duct tumor, Malignant cystic duct tumor, Malignant common bile duct tumor, Malignant extrahepatic bile duct tumor, Malignant hepatobiliary tumor, Malignant hilar bile duct tumor, Common bile duct cancer, Common bile duct CA, Lower common bile duct cancer, Lower common bile duct CA, Common bile duct adenocarcinoma, Cholangiocarcinoma, Bile duct cell carcinoma, Lower bile duct cancer, Hepatic duct cancer, Malignant hepatic duct tumor. |

| | | |
| --- | --- | --- |
| Gallbladder Carcinoma | C23 | Gallbladder cancer, Gallbladder adenocarcinoma, Gallbladder cystadenocarcinoma, Gallbladder squamous cell carcinoma, Gallbladder cystic squamous cell carcinoma, Gallbladder CA, Malignant gallbladder tumor. |
| Urologic Neoplasms | | |
| Renal Carcinoma | C64, C65 | Kidney cancer, Renal cell carcinoma, RCC, Renal parenchymal epithelial tumor, Malignant kidney tumor, Clear cell carcinoma of the kidney, Kidney CA, Secondary malignant kidney tumor, Malignant renal tumor, Kidney tumor, Malignant |
| Bladder Cancer | C67 | Bladder cancer, Malignant bladder tumor, Bladder CA, Malignant tumor of the bladder dome, Malignant tumor of the bladder neck, Malignant tumor of the anterior wall of the bladder, Malignant tumor of the bladder trigone, Transitional cell carcinoma of the bladder, Malignant tumor of the posterior wall of the bladder, Malignant tumor crossing the bladder neck, Malignant tumor of the lateral wall of the bladder, Bladder adenocarcinoma, Malignant tumor of the ureteral orifice, Urachal carcinoma, Multiple malignant tumors of the bladder. |
| Ureteral Carcinoma | C66 | Ureteral cancer, Malignant ureteral tumor, Ureteral CA. |
| Female Reproductive Carcinoma | | |
| Cervical Cancer | C53 | Cervical cancer, Malignant cervical tumor, Cervical CA, Cervical adenocarcinoma, Cervical squamous cell carcinoma, Well-differentiated cervical adenocarcinoma, Poorly differentiated squamous cell carcinoma of the cervix, Moderately differentiated squamous carcinoma, Moderately differentiated squamous cell carcinoma of the cervix, Non-keratinizing squamous cell carcinoma of the cervix. |

| | | |
|---|---|---|
| Endometrial Cancer | C54, C55 | Uterine cancer, Malignant endometrial tumor, Endometrial CA, Endometrial wall carcinoma, Endometrial adenocarcinoma, Endometrioid adenocarcinoma, Moderately differentiated endometrial adenocarcinoma, Well-differentiated endometrial adenocarcinoma, Mixed endometrial carcinoma, Serous endometrial adenocarcinoma, Endometrioid carcinoma, Endometrial stromal sarcoma, Serous carcinoma of the endometrium, Poorly differentiated endometrial adenocarcinoma, High-grade serous endometrial carcinoma, Clear cell carcinoma of the endometrium, Moderately differentiated endometrioid adenocarcinoma, Uterine body cancer, Malignant uterine tumor, Uterine body CA, Endometrial epithelial carcinoma. |
| Ovarian Cancer | C56 | Ovarian cancer, Malignant ovarian tumor, Ovarian CA, Recurrent malignant ovarian tumor, High-grade serous ovarian cancer, High-grade serous papillary adenocarcinoma of the ovary, Poorly differentiated serous adenocarcinoma of the ovary, Secondary malignant ovarian tumor, Serous adenocarcinoma of the ovary, Serous cystadenocarcinoma of the ovary, Granulosa cell tumor of the ovary, Papillary cystadenocarcinoma of the ovary, Clear cell carcinoma of the ovary, Ovarian adenocarcinoma, Mucinous adenocarcinoma of the ovary, Small cell carcinoma of the ovary |
| Hematologic Neoplasms | | |
| Leukemia | C91-C96 | Leukemia, Blood cancer, Chronic lymphocytic leukemia (CLL), Acute myeloid leukemia (AML). |
| Lymphoma | C81-C88 | Lymphoma, Malignant lymphoma, (Non-)Hodgkin lymphoma, Diffuse large B-cell lymphoma. |
| Myeloma | C90.0 | Multiple myeloma. |
| Others System | | |

| | | |
|---|---|---|
| Lung Cancer | C34 | Lung cancer, Lung Ca, RLC, LLC, Malignant lung tumor, Poorly differentiated lung cancer, Lung squamous cell carcinoma, Lung squamous cell carcinoma, Lung non-small cell carcinoma, Lung adenocarcinoma, Lung small cell carcinoma, Central type lung squamous cell carcinoma, Poorly differentiated squamous cell carcinoma of the lower lobe of the lung, Lung small cell carcinoma, Recurrent malignant lung tumor, Alveolar cell carcinoma, Multiple metastatic lung cancer, Poorly differentiated adenocarcinoma of the lung, Malignant bronchial tumor, Alveolar adenocarcinoma, Lung tumor, Malignant |
| Thyroid Cancer | C73 | Thyroid cancer, Malignant thyroid tumor, Thyroid CA, Thyroid carcinoma, Thyroid carcinoma, Multiple malignant thyroid tumors, Papillary thyroid carcinoma, Thyroid adenocarcinoma, Malignant thyroid tumor. |
| Breast Cancer | C50 | Breast cancer, Malignant breast tumor, BC, Malignant tumor of the breast, Breast carcinoma, Breast CA, Mammary CA, Invasive ductal carcinoma, Invasive ductal carcinoma of the breast, Invasive breast cancer, Malignant tumor of the mammary gland, Ductal carcinoma of the breast, Malignant tumor of the upper inner quadrant of the breast, Malignant breast tumor, Breast tumor, Malignant |
| Prostate Cancer | C61 | Prostate cancer, PC, Malignant prostate tumor, Prostate CA, Poorly differentiated prostate adenocarcinoma, Recurrent malignant prostate tumor, Prostate adenocarcinoma, Moderately differentiated prostate cancer, Prostate metastatic cancer, Prostate tumor, Malignant |
| Oropharyngeal Cancer | C00-C10 | Palate cancer, Nasal cancer, Pharyngeal cancer, Malignant nasal tumor, Malignant pharyngeal tumor, Nasal CA, Pharyngeal CA, Buccal cancer, Tongue cancer, Gingival cancer, Floor of mouth cancer, Oral cancer, Malignant palate tumor, Malignant buccal tumor, Malignant tongue tumor, Malignant gingival tumor, Malignant floor of mouth tumor, Malignant oral tumor, Palate CA, Buccal CA, Tongue CA, Gingival CA, Floor of mouth CA, Oral CA. |

| | | |
|---|---|---|
| Laryngeal Cancer | C32 | Laryngeal cancer, Malignant laryngeal tumor, Laryngeal CA, Supraglottic cancer, Glottic cancer, Subglottic cancer, Vocal cord cancer, Malignant supraglottic tumor, Malignant glottic tumor, Malignant subglottic tumor, Malignant vocal cord tumor. |

**Ascertainment of pharmacological treatments of the metabolic dysfunction**

We identified agents primarily through their action receptors or pharmacological classification and searched our database for prescriptions using relevant keywords, as detailed in supplementary table 2. Furthermore, various combination medications are utilized in clinical practice. Therefore, we manually categorized them in our study.

**Included comorbidities**

As indicated by previous studies on the natural history of NAFLD or MASLD, various comorbidities coexist or interact bidirectionally with cardiometabolic factors(1). These include cardiovascular disease (CVD), abnormal liver function, viral hepatitis, cirrhosis, chronic kidney disease (CKD), obstructive sleep apnea (OSA), polycystic ovarian syndrome (PCOS), H. pylori infection, autoimmune hepatitis, and chronic obstructive pulmonary disease (COPD). Due to the leakage of smoking data in our database, COPD has been considered a proxy for smoking in this study(2)

Supplementary Table 2. Anti-metabolic dysfunction agents

| Keywords and forms | Name |
|---|---|
| Hyperglycemia | |
| Insulin | |
| | Insulin degludec and insulin aspart |
| | Insulin glargine |
| | Insulin aspart |
| | Insulin aspart 30 |
| | Insulin aspart 50 |
| | Insulin degludec/liraglutide |
| Metformin | |
| Sodium-Glucose Cotransporter-2 Inhibitors | |
| | Ertugliflozin |
| | Dapagliflozin |
| | Empagliflozin |
| | Canagliflozin |
| GLP-1 Receptor Agonists | |
| | Exenatide |
| | Brezapride |
| | Dulaglutide |
| | Pegylated Liraglutide |
| | Liraglutide |
| | Lixisenatide |
| | Semaglutide |
| | Insulin degludec/liraglutide |
| Dipeptidyl Peptidase-4 Inhibitors | |
| | Alogliptin |
| | Linagliptin |
| | Saxagliptin |
| | Sitagliptin |
| | Vildagliptin |
| | Sitagliptin |
| PPAR-γ agonists (Thiazolidinediones) | |
| | Pioglitazone |
| | Rosiglitazone |
| Dyslipidemia | |
| Statins | |
| | Atorvastatin |
| Fibrates | |
| | Fenofibrate |
| | Benzafibrate |
| Cholesterol Absorption Inhibitors | |
| | Ezetimibe |
| Hypertension | |

| | |
|---|---|
| Calcium Channel Blockers | |
| | Amlodipine |
| | Benidipine |
| | Felodipine |
| | Lacidipine |
| | Lercanidipine |
| | Nicardipine |
| | Nimodipine |
| | Nisoldipine |
| | Sildenafil |
| | Nifedipine |
| | Nifedipine |
| | L-amlodipine |
| ACEIs/ARBs | |
| | Aliskiren |
| | Olmesartan Medoxomil |
| | Benazepril |
| | Irbesartan |
| | Fosinopril |
| | Captopril |
| | Candesartan Cilexetil |
| | Lisinopril |
| | Ramipril |
| | Losartan |
| | Midapril |
| | Perindopril |
| | Telmisartan |
| | Valsartan |
| | Enalapril |
| Beta-Blockers | |
| | Arolol |
| | Atenolol |
| | Esmolol |
| | Bisoprolol |
| | Labetalol |
| | Landiolol |
| | Metoprolol |
| | Nipradilol |
| | Propranolol |
| | Timolol |
| | Sotalol |

ACEIs/ARBs:Angiotensin-Converting Enzyme Inhibitors/Angiotensin II Receptor Blockers

Supplementary Table 3a. Basic clinical characteristic of MASLD with extrahepatic cancers

| | N (%) |
|---|---|
| Total | 6,605 |
| Gender, female | 3,243(49.1) |
| Age, year | |
| <30 | 105(1.59) |
| 30-39 | 380(5.75) |
| 40-49 | 913(13.82) |
| 50-59 | 2,147(32.51) |
| 60-69 | 1,839(27.84) |
| ≥70 | 1,221(18.49) |
| Cardiometabolic factors | |
| Hypertension | 3,271(49.52) |
| Blood pressure ≥130/85mmHg | 3,034(45.93) |
| Utility of Antihypertensive drug | 1,183(17.91) |
| Dyslipidemia | 4,275(64.72) |
| Diagnosis of Hyperlipidemia | 1,666(25.22) |
| Triglycerides≥1.7mmol/L | 2,401(36.35) |
| Lower HDL-C | 2,568(38.88) |
| LDL-C ≥3.4 mmol/L | 1,143(17.31) |
| Total Cholesterol ≥5.2 mmol/L | 1,420(21.5) |
| Utility of Lipid-lowering Agent | 738(11.17) |
| Hyperglycemia | 3,237(49.01) |
| Type 2 Diabetes Mellitus | 2,691(40.74) |
| Fasting Blood-glucose 5.6-6.9mmol/L | 629(9.52) |
| HbA1c 5.7-6.4% | 810(12.26) |
| 2-hour post-meal blood glucose 7.8-11.0mmol/L | 38(0.58) |
| Utility of hypoglycemic agent | 903(13.67) |
| Other Related Indicators | |
| HOMA-IR≥2.5 | 13(0.2) |
| hs-CRP over 2mg/L | 1281(19.39) |
| Combination of cardiometabolic risk factors | |
| Hypertension and abnormal lipid metabolism | 1,760(26.65) |
| Hypertension and hyperglycemia | 1,570(23.77) |
| Abnormal lipid metabolism and hyperglycemia | 1,857(28.12) |
| Hypertension, abnormal lipid metabolism and hyperglycemia | 1,009(15.28) |
| Liver function | |
| AST>40U/L | 1,097(16.61) |
| AST>80U/L | 333(5.04) |
| ALT>40U/L | 1,608(24.35) |
| ALT>80U/L | 522(7.9) |
| Fibrosis-4 index | |
| FIB-4<1.3 | 2,274(34.43) |

| | |
|---|---|
| FIB-4>1.3 | 3,169(47.98) |
| FIB-4 1.3-2.66 | 2,190(33.16) |
| FIB-4≥2.67 | 1,011(15.31) |
| FIB-4≥3.48 | 649(9.83) |

ALT, alanine aminotransferase, AST, aspartate aminotransferase, HDL-C: high density lipoprotein cholesterol, LDL-C: low-density lipoprotein cholesterol, HOMA-IR: insulin resistance index, FIB-4: Fibrosis-4 index, hs-CRP: high-sensitivity c-reactive protein

Supplementary Table 3b. Basic comorbidities and metabolic-based treatments of MASLD with extrahepatic cancers

| Comorbidities | |
|---|---|
| Cardiovascular Disease | 3445(52.16) |
| Abnormal Liver Function | 509(7.71) |
| Viral Hepatitis | 233(3.53) |
| Cirrhosis | 50(0.76) |
| Chronic Kidney Disease | 258(3.91) |
| Osteoporosis | 307(4.65) |
| Hypothyroidism | 302(4.57) |
| Obstructive Sleep Apnea | 41(0.62) |
| Polycystic Ovarian Syndrome | 6(0.09) |
| Hp. Infection | 83(1.26) |
| Autoimmune hepatitis | 7(0.11) |
| Chronic obstructive pulmonary disease | 134(2.03) |
| Metabolic-based treatments | |
| Hyperglycemia | |
| Insulin (Aspart Insulin and Glargine Insulin) | 226(3.42) |
| Biguanides(Metformin) | 404(6.12) |
| SGLT-2 Inhibitors | 87(1.32) |
| GLP-1 Receptor Agonists | 28(0.42) |
| DPP-4 Inhibitors | 101(1.53) |
| Thiazolidinediones | 13(0.2) |
| Dyslipidemia | |
| Statins | 442(6.69) |
| Fibrates | 47(0.71) |
| Cholesterol Absorption Inhibitors (Ezetimibe) | 37(0.56) |
| Hypertension | |
| Calcium Channel Blockers | 705(10.67) |
| ACEIs/ARBs | 451(6.83) |
| Beta-Blockers | 282(4.27) |

SGLT-2: Sodium-Glucose Cotransporter-2; GLP-1: Glucagon-like peptide-1; DPP-4: Dipeptidyl Peptidase-4; ACEIs/ARBs: Angiotensin-Converting Enzyme Inhibitors/Angiotensin II Receptor Blockers.

Supplementary Table 4. Gender difference of site-specific extrahepatic cancers in MASLD population

| Cancer category | Total | Male (%) | Female (%) | *P* |
|---|---|---|---|---|
| Extrahepatic Cancers | 6,605 | 3362(50.9) | 3243(49.1) | 0.1431 |
| Digestive System | 1,699 | 1102(64.86) | 597(35.14) | <0.001 |
| Colorectal Carcinoma | 892 | 570(63.9) | 322(36.1) | <0.001 |
| Gastric Carcinoma | 445 | 310(69.66) | 135(30.34) | <0.001 |
| Pancreatic Carcinoma | 155 | 89(57.42) | 66(42.58) | 0.0647 |
| Esophageal Carcinoma | 127 | 101(79.53) | 26(20.47) | <0.001 |
| Extrahepatic Cholangiocarcinoma | 102 | 57(55.88) | 45(44.12) | 0.2348 |
| Gallbladder Carcinoma | 62 | 28(45.16) | 34(54.84) | 0.4461 |
| Urinary System | 693 | 510(73.59) | 183(26.41) | <0.001 |
| Renal Carcinoma | 510 | 367(71.96) | 143(28.04) | <0.001 |
| Bladder Cancer | 170 | 137(80.59) | 33(19.41) | <0.001 |
| Ureteral Carcinoma | 26 | 14(53.85) | 12(46.15) | 0.6949 |
| Female Reproductive System | 631 | | 631(100) | |
| Cervical Cancer | 260 | | 260(100) | |
| Endometrial Cancer | 257 | | 257(100) | |
| Ovarian Cancer | 162 | | 162(100) | |
| Hematological System | 815 | 483(59.26) | 332(40.74) | <0.001 |
| Leukemia | 292 | 179(61.3) | 113(38.7) | <0.001 |
| Lymphoma | 427 | 253(59.25) | 174(40.75) | <0.001 |
| Myeloma | 144 | 83(57.64) | 61(42.36) | 0.0668 |
| Others Systems | 4,623 | 2265(48.99) | 2358(51.01) | 0.1714 |
| Lung Cancer | 2,191 | 1251(57.1) | 940(42.9) | <0.001 |
| Thyroid Cancer | 1,186 | 593(50) | 593(50) | 0.9077 |
| Breast Cancer | 924 | 15(1.62) | 909(98.38) | <0.001 |
| Prostate Cancer | 298 | 298(100) | | |
| Oropharyngeal Cancer | 105 | 83(79.05) | 22(20.95) | <0.001 |
| Laryngeal Cancer | 70 | 67(95.71) | 3(4.29) | <0.001 |

Supplementary Table 5. MASLD with extrahepatic cancers risk factors. Univariate logistic regression analysis

| | Extrahepatic cancers | | OR (95%CI) | *P* |
|---|---|---|---|---|
| | Yes | No | | |
| Age | | | | |
| <30 | 105 | 5,979 | Reference | |
| 30-39 | 380 | 16,111 | 1.34(1.08,1.67) | 0.0080 |
| 40-49 | 913 | 20,855 | 2.49(2.03,3.06) | <0.001 |
| 50-59 | 2,147 | 27,536 | 4.44(3.64,5.41) | <0.001 |
| 60-69 | 1,839 | 16,619 | 6.30(5.16,7.69) | <0.001 |
| >70 | 1,221 | 9,947 | 6.99(5.71,8.55) | <0.001 |
| Gender, female | 3,362 | 64,059 | 1.87(1.78,1.79) | <0.001 |
| Cardiometabolic factors | | | | |
| Hypertension | | | | |
| Blood pressure ≥130/85mmHg | 3,034 | 38,220 | 1.31(1.24,1.37) | <0.001 |
| Utility of Antihypertensive drug | 1,183 | 23,036 | 0.70(0.66,0.75) | <0.001 |
| Dyslipidemia | | | | |
| Diagnosis of Hyperlipidemia | 1,666 | 40,069 | 0.48(0.45,0.51) | <0.001 |
| Triglycerides over 1.7mmol/L | 2,568 | 39,317 | 0.93(0.89,0.98) | 0.0089 |
| Lower HDL-C | 1,420 | 25,029 | 0.79(0.74,0.84) | <0.001 |
| LDL-C over 3.4 mmol/L | 1,143 | 21,308 | 0.74(0.70,0.79) | <0.001 |
| Total Cholesterol over 5.2 mmol/L | 2,401 | 44,356 | 0.68(0.64,0.71) | <0.001 |
| Utility of Lipid-lowering Agent | 738 | 23,679 | 0.39(0.36,0.42) | <0.001 |
| Hyperglycemia | | | | |
| Type 2 Diabetes Mellitus | 2,691 | 36,312 | 1.15(1.09,1.21) | <0.001 |
| pre-diabetes | 1,349 | 20,162 | 0.98(0.92,1.04) | 0.4978 |
| Utility of hypoglycemic agent | 903 | 19,231 | 0.64(0.60,0.69) | <0.001 |
| Other Related Indicators | | | | |
| HOMA-IR≥2.5 | 13 | 974 | 0.19(0.11,0.34) | <0.001 |
| Hs-CRP over 2mg/L | 1,281 | 9,175 | 2.30(2.16,2.46) | <0.001 |
| Cardiometabolic factors' classification | | | | |
| Hypertension | 3,271 | 40,446 | 1.37(1.31,1.44) | <0.001 |
| Abnormal lipid metabolism | 4,275 | 80,213 | 0.38(0.37,0.41) | <0.001 |
| Hyperglycemia | 3,237 | 45,109 | 1.11(1.05,1.16) | <0.001 |
| Combination of cardiometabolic dysfunction | | | | |
| Hypertension and abnormal lipid metabolism | 1,760 | 31,287 | 0.76(0.72,0.81) | <0.001 |
| Hypertension and hyperglycemia | 1,570 | 22,047 | 1.06(1.00,1.12) | 0.0486 |
| Abnormal lipid metabolism and hyperglycemia | 1,857 | 34,006 | 0.73(0.69,0.77) | <0.001 |
| Hypertension, abnormal lipid metabolism and hyperglycemia | 1,009 | 18,619 | 0.76(0.71,0.81) | <0.001 |
| Liver function | | | | |
| AST>40U/L | 1,097 | 15,199 | 1.07(1.00,1.15) | 0.0408 |

| | | | | |
|---|---|---|---|---|
| AST>80U/L | 333 | 4,497 | 1.09(0.97,1.22) | 0.1282 |
| ALT>40U/L | 1,608 | 26,301 | 0.87(0.82,0.92) | <0.001 |
| ALT>80U/L | 522 | 9,191 | 0.82(0.75,0.90) | <0.001 |
| Comorbidities | | | | |
| Cardiovascular Disease | 3,445 | 44,816 | 1.27(1.21,1.34) | <0.001 |
| Abnormal Liver Function | 509 | 10,340 | 0.70(0.64,0.77) | <0.001 |
| Viral Hepatitis | 233 | 9,526 | 0.34(0.29,0.38) | <0.001 |
| Cirrhosis | 50 | 2,011 | 0.36(0.27,0.48) | <0.001 |
| Chronic Kidney Disease | 258 | 7,388 | 0.49(0.43,0.56) | <0.001 |
| Obstructive Sleep Apnea | 41 | 1,516 | 0.39(0.29,0.54) | <0.001 |
| Polycystic Ovarian Syndrome | 6 | 319 | 0.28(0.12,0.62) | 0.0018 |
| Hp. Infection | 83 | 1,805 | 0.67(0.54,0.84) | <0.001 |
| Autoimmune hepatitis | 7 | 297 | 0.35(0.16,0.73) | 0.0055 |
| Chronic obstructive pulmonary disease | 134 | 874 | 2.28(1.90,2.74) | <0.001 |
| Liver fibrosis FIB-4 index | | | | |
| Age 35-65 | | | | |
| FIB-4<1.3 as reference | 1,789 | 28,282 | Reference | |
| FIB-4 1.3-2.66 | 1,175 | 12,323 | 1.51(1.40,1.63) | <0.001 |
| FIB-4≥2.67 | 130 | 1,159 | 1.77(1.47,2.14) | <0.001 |
| FIB-4≥3.48 | 267 | 2,216 | 1.90(1.66,2.18) | <0.001 |
| FIB-4 1.3-2.66 as reference | 1,175 | 12,323 | Reference | |
| FIB-4<1.3 | 1,789 | 28,282 | 0.66(0.61,0.72) | <0.001 |
| FIB-4≥2.67 | 130 | 1,159 | 1.18(0.97,1.42) | 0.0954 |
| FIB-4≥3.48 | 267 | 2,216 | 1.26(1.10,1.45) | 0.0011 |
| Age over 65 | | | | |
| FIB-4<2 | 934 | 7,696 | Reference | |
| FIB-4≥2 | 955 | 5,975 | 1.32(1.20,1.45) | <0.001 |

T2DM: type 2 diabetes mellitus, HDL-C: High density lipoprotein cholesterol, LDL-C: Low-Density Lipoprotein cholesterol, HOMA-IR: insulin resistance index, FIB-4: Fibrosis-4 index, hsCRP: High-sensitivity C-reactive protein. ALT, Alanine Aminotransferase, AST, Aspartate Aminotransferase

Supplementary Table 6. Hypertension subgroup the risk factors of extrahepatic cancers. Univariate logistic regression analyses

| Covariate | Extrahepatic cancers | | OR(95%CI) | *P* |
|---|---|---|---|---|
| | Yes | No | | |
| Age | | | | |
| <30 | 8 | 941 | Reference | |
| 30-34 | 34 | 1,405 | 2.85(1.31,6.18) | 0.0081 |
| 35-39 | 49 | 2,149 | 2.68(1.27,5.68) | 0.0101 |
| 40-44 | 92 | 2,637 | 4.10(1.98,8.48) | <0.001 |
| 45-49 | 216 | 4,107 | 6.19(3.04,12.57) | <0.001 |
| 50-54 | 390 | 5,661 | 8.10(4.01,16.37) | <0.001 |
| 55-59 | 582 | 6,732 | 10.17(5.04,20.50) | <0.001 |
| 60-64 | 441 | 4,522 | 11.47(5.68,23.16) | <0.001 |
| 65-70 | 610 | 5,207 | 13.78(6.84,27.77) | <0.001 |
| ≥70 | 849 | 7,085 | 14.09(7.00,28.37) | <0.001 |
| Female gender | 1,532 | 14,722 | 1.54(1.43,1.65) | <0.001 |
| Hyperglycemia | 1,570 | 22,047 | 0.77(0.72,0.83) | <0.001 |
| Abnormal lipid metabolism | 1,760 | 31,287 | 0.34(0.32,0.37) | <0.001 |
| T2DM | 1,361 | 18,168 | 0.87(0.81,0.94) | <0.001 |
| Using hypoglycemia treatment | 516 | 10,213 | 0.55(0.50,0.61) | <0.001 |
| Hyperlipidemia | 744 | 17,543 | 0.38(0.35,0.42) | <0.001 |
| Using adjust lipid agents | 466 | 13,704 | 0.32(0.29,0.36) | <0.001 |
| Abnormal liver function | 194 | 2,968 | 0.80(0.69,0.92) | 0.0029 |
| Cirrhosis | 22 | 449 | 0.60(0.39,0.93) | 0.0211 |
| Chronic kidney disease | 189 | 5,264 | 0.41(0.35,0.48) | <0.001 |
| Cardiovascular disease | 3,080 | 38,832 | 0.67(0.57,0.78) | <0.001 |
| Obstructive Sleep Apnea | 33 | 1,041 | 0.39(0.27,0.55) | <0.001 |
| Hypothyroidism | 158 | 1,404 | 1.41(1.19,1.67) | <0.001 |
| Viral Hepatitis | 98 | 1,624 | 0.74(0.60,0.91) | 0.0041 |
| Hp. Infection | 35 | 744 | 0.58(0.41,0.81) | 0.0016 |
| Chronic obstructive pulmonary disease | 84 | 560 | 1.88(1.49,2.37) | <0.001 |
| AST>40U/L | 477 | 4,909 | 1.24(1.12,1.37) | <0.001 |
| Liver fibrosis FIB-4 index | | | | |
| Age 35-65 | | | | |
| FIB-4<1.3 | 746 | 11,529 | Reference | |
| FIB-4 1.3-2.66 | 533 | 5,354 | 1.54(1.37,1.73) | <0.001 |
| FIB-4≥2.67 | 50 | 510 | 1.52(1.12,2.04) | 0.0066 |
| FIB-4≥3.48 | 92 | 834 | 1.70(1.36,2.14) | <0.001 |
| FIB-4 1.3-2.66 | 533 | 5,354 | Reference | |
| FIB-4<1.3 | 746 | 11,529 | 0.65(0.58,0.73) | <0.001 |
| FIB-4≥2.67 | 50 | 510 | 0.95(0.73,1.33) | 0.9213 |
| FIB-4≥3.48 | 92 | 834 | 1.11(0.88,1.40) | 0.3879 |
| Over 65-year-old | | | | |

| | | | | |
|---|---|---|---|---|
| FIB-4<2 | 603 | 5,349 | Reference | |
| FIB-4≥2 | 607 | 4,109 | 1.31(1.16,1.48) | <0.001 |

Supplementary Table 7. Abnormal lipid metabolism subgroup the risk factors of extrahepatic cancers. Univariate logistic regression analyses

| Covariate | Extrahepatic cancers | | OR (95%CI) | *P* |
|---|---|---|---|---|
| | YES | NO | | |
| Age | | | | |
| <30 | 88 | 5,137 | Reference | |
| 30-34 | 118 | 6,524 | 1.06(0.80,1.39) | 0.7022 |
| 35-39 | 164 | 7,426 | 1.29(0.99,1.67) | 0.0569 |
| 40-44 | 249 | 7,672 | 1.89(1.48,2.42) | <0.001 |
| 45-49 | 407 | 9,882 | 2.40(1.90,3.03) | <0.001 |
| 50-54 | 594 | 11,029 | 3.14(2.51,3.94) | <0.001 |
| 55-59 | 767 | 11,210 | 3.99(3.20,4.99) | <0.001 |
| 60-64 | 505 | 6,525 | 4.52(3.59,5.68) | <0.001 |
| 65-70 | 593 | 6,809 | 5.08(4.05,6.38) | <0.001 |
| ≥70 | 790 | 7,999 | 5.77(4.61,7.21) | <0.001 |
| Female | 2,186 | 26,702 | 2.10(1.97,2.23) | <0.001 |
| Hyperglycemia | 1,857 | 34,006 | 1.04(0.98,1.11) | 0.1785 |
| Hypertension | 1,760 | 31,287 | 1.09(1.03,1.16) | 0.0047 |
| T2DM | 1,515 | 27,033 | 1.08(1.01,1.15) | 0.0193 |
| Using hypoglycemia treatment | 646 | 16,147 | 0.71(0.65,0.77) | <0.001 |
| pre-diabetes | 514 | 9,956 | 0.96(0.88,1.06) | 0.4580 |
| Hypertension(diagnosis) | 1,659 | 29,840 | 1.07(1.01,1.14) | 0.0344 |
| Using hypertension treatment | 841 | 19,383 | 0.77(0.71,0.83) | <0.001 |
| Hyperglycemia and Hypertension | 1,009 | 18,619 | 1.02(0.95,1.10) | 0.5538 |
| Abnormal liver function | 350 | 8,693 | 0.73(0.66,0.82) | <0.001 |
| Cirrhosis | 37 | 1,617 | 0.42(0.31,0.59) | <0.001 |
| Chronic kidney disease | 204 | 6,408 | 0.58(0.50,0.67) | <0.001 |
| Cardiovascular disease | 1,985 | 35,775 | 1.08(1.01,1.15) | 0.0189 |
| Osteoporosis | 247 | 3,554 | 1.32(1.16,1.51) | <0.001 |
| Obstructive Sleep Apnea | 33 | 1,319 | 0.47(0.33,0.66) | <0.001 |
| Polycystic Ovarian Syndrome | 4 | 228 | 0.33(0.12,0.88) | 0.0276 |
| Hypothyroidism | 247 | 2,368 | 2.02(1.76,2.31) | <0.001 |
| Viral Hepatitis | 147 | 8,178 | 0.31(0.27,0.37) | <0.001 |
| Chronic obstructive pulmonary disease | 89 | 672 | 2.52(2.01,3.15) | <0.001 |
| AST>40U/L | 731 | 12,799 | 1.09(1.00,1.18) | 0.0471 |
| ALT>40U/L | 1,100 | 22,653 | 0.88(0.82,0.94) | 0.0004 |
| Liver fibrosis FIB-4 index | | | | |
| Age 35-65 | | | | |

| | | | | |
|---|---|---|---|---|
| FIB-4<1.3 | 1,261 | 24,581 | Reference | |
| FIB-4 1.3-2.66 | 752 | 10,366 | 1.55(1.29,1.55) | <0.001 |
| FIB-4≥2.67 | 91 | 935 | 1.90(1.52,2.37) | <0.001 |
| FIB-4≥3.48 | 174 | 1,782 | 1.90(1.61,2.25) | <0.001 |
| FIB-4 1.3-2.66 | 752 | 10,366 | Reference | |
| FIB-4<1.3 | 1,261 | 24,581 | 0.71(0.64,0.78) | <0.001 |
| FIB-4≥2.67 | 91 | 935 | 1.34(1.07,1.68) | 0.0114 |
| FIB-4≥3.48 | 174 | 1,782 | 1.35(1.13,1.60) | <0.001 |
| Over 65-year-old | | | | |
| FIB-4<2 | 588 | 6,663 | Reference | |
| FIB-4≥2 | 612 | 4,969 | 1.40(1.24,1.57) | <0.001 |

Supplementary Table 8. T2DM subgroup the risk factors of extrahepatic cancers. Univariate logistic regression analyses

| Covariate | Extrahepatic cancers | | OR (95%CI) | *P* |
|---|---|---|---|---|
| | YES | NO | | |
| Age | | | | |
| <30 | 34 | 1,661 | Reference | |
| 30-34 | 54 | 1,885 | 1.40(0.91,2.16) | 0.1292 |
| 35-39 | 61 | 2,526 | 1.18(0.77,1.80) | 0.4449 |
| 40-44 | 112 | 2,797 | 1.96(1.33,2.89) | <0.001 |
| 45-49 | 212 | 3,996 | 2.59(1.80,3.74) | <0.001 |
| 50-54 | 324 | 5,175 | 3.06(2.14,4.37) | <0.001 |
| 55-59 | 501 | 5,925 | 4.13(2.91,5.87) | <0.001 |
| 60-64 | 355 | 3,873 | 4.48(3.13,6.40) | <0.001 |
| 65-70 | 486 | 4,131 | 5.75(4.04,8.17) | <0.001 |
| ≥70 | 599 | 5,412 | 5.41(3.81,7.67) | <0.001 |
| Female | 1,267 | 13,612 | 1.50(1.39,1.63) | 0.0000 |
| Hypertension | 1,383 | 18,633 | 1.03(0.95,1.11 | 0.5016 |
| Abnormal lipid metabolism | 1,552 | 27,947 | 0.44(0.41,0.48) | <0.001 |
| Hypertension diagnoses | 1,312 | 17,831 | 1.01(0.93,1.09) | 0.8259 |
| Using hypertension treatment | 592 | 10,959 | 0.67(0.61,0.73) | <0.001 |
| Hyperlipidemia | 703 | 16,830 | 0.42(0.39,0.46) | <0.001 |
| Using adjust lipid agents | 360 | 10,493 | 0.39(0.35,0.43) | <0.001 |
| Hypertension and abnormal lipid metabolism | 876 | 15,638 | 0.65(0.60,0.71) | <0.001 |
| Abnormal liver function | 228 | 3,861 | 0.79(0.69,0.91) | <0.001 |
| Cirrhosis | 22 | 620 | 0.48(0.31,0.74) | <0.001 |
| Chronic kidney disease | 180 | 4,612 | 0.50(0.43,0.58) | <0.001 |
| Obstructive Sleep Apnea | 17 | 601 | 0.38(0.24,0.62) | <0.001 |
| Polycystic Ovarian Syndrome | 5 | 221 | 0.31(0.13,0.75) | 0.0093 |

| | | | | |
|---|---|---|---|---|
| Hypothyroidism | 133 | 1,251 | 1.47(1.23,1.77) | <0.001 |
| Viral Hepatitis | 97 | 2,081 | 0.62(0.51,0.77) | <0.001 |
| Hp. Infection | 21 | 711 | 0.40(0.26,0.62) | <0.001 |
| Chronic obstructive pulmonary disease | 56 | 437 | 1.77(1.33,2.34) | <0.001 |
| AST>40U/L | 473 | 5,892 | 1.12(1.01,1.24) | 0.0365 |
| Liver fibrosis FIB-4 index | | | | |
| Age 35-65 | | | | |
| FIB-4<1.3 | 578 | 9,584 | Reference | |
| FIB-4 1.3-2.66 | 459 | 4,914 | 1.55(1.36,1.76) | <0.001 |
| FIB-4≥2.67 | 52 | 529 | 1.63(1.21,2.19) | 0.0013 |
| FIB-4≥3.48 | 135 | 1,090 | 2.05(1.69,2.50) | <0.001 |
| FIB-4 1.3-2.66 | 459 | 4,914 | Reference | |
| FIB-4<1.3 | 578 | 9,584 | 0.65(0.57,0.73) | <0.001 |
| FIB-4≥2.67 | 52 | 529 | 1.05(0.78,1.42) | 0.7391 |
| FIB-4≥3.48 | 135 | 1,090 | 1.33(1.08,1.62) | 0.0064 |
| Over 65-year-old | | | | |
| FIB-4<2 | 438 | 3,981 | Reference | |
| FIB-4≥2 | 452 | 3,030 | 1.36(1.18,1.56) | <0.001 |

**Ascertainment of adjusted covariate in each subgroup**

The adjusted covariate in each subgroup during analysis on **Pharmacological treatments of the metabolic dysfunction showed protective impaction on extrahepatic cancers in MASLD population** as following:

Hypertension subgroup adjusted factors: age, sex, hyperglycemia, abnormal lipid metabolism, T2DM, using hypoglycemia treatment, hyperlipidemia, using adjust lipid agents, abnormal liver function, cirrhosis, chronic kidney disease, cardiovascular disease, obstructive sleep apnea, hypothyroidism, viral hepatitis, hp. infection, chronic obstructive pulmonary disease, AST>40u/l

Abnormal lipide metabolism group adjusted factors: age, sex, hyperglycemia, hypertension, T2DM, using hypoglycemia treatment, pre-diabetes, hypertension(diagnosis), using hypertension treatment, hyperglycemia and hypertension, abnormal liver function, cirrhosis, chronic kidney disease, cardiovascular disease, osteoporosis, obstructive sleep apnea, polycystic ovarian syndrome, hypothyroidism, viral hepatitis, chronic obstructive pulmonary disease, AST>40U/L, ALT>40U/L

T2DM subgroup adjusted factors: age, sex, hypertension, abnormal lipid metabolism, hypertension diagnoses, using hypertension treatment, hyperlipidemia, using adjust lipid agents, hypertension and abnormal lipid metabolism, abnormal liver function, cirrhosis, chronic kidney disease, obstructive sleep apnea, polycystic ovarian syndrome, hypothyroidism, viral hepatitis, hp. infection, chronic obstructive pulmonary disease, AST>40U/L

The adjusted covariate in each subgroup during analysis on **Fibrosis 4 score associated with extrahepatic cancers in MASLD population** as following:

All MASLD population adjusted factors: Gender, hypertension, hyperlipidemia, type 2 diabetes mellitus, pre-diabetes, anti-metabolic dysfunction agents, abnormal blood pressure, abnormal lipid metabolism, hyperglycemia, the combination of metabolic dysfunction, hs-CRP, cardiovascular disease, cirrhosis, chronic obstructive pulmonary disease

Hypertension subgroup adjusted factors: sex, Hyperglycemia, Abnormal lipid metabolism, T2DM, Using hypoglycemia treatment, Hyperlipidemia, Using adjust lipid agents, Abnormal liver function, Cirrhosis, Chronic kidney disease, Cardiovascular disease, Obstructive Sleep Apnea, Hypothyroidism, Viral Hepatitis, Hp. Infection, Chronic obstructive pulmonary disease

Abnormal lipide metabolism group adjusted factors: sex, Hyperglycemia, Hypertension, T2DM, Using hypoglycemia treatment, pre-diabetes, Hypertension(diagnosis), Using hypertension treatment, Hyperglycemia and Hypertension, Abnormal liver function, Cirrhosis, Chronic kidney disease, Cardiovascular disease, Osteoporosis, Obstructive Sleep Apnea, Polycystic Ovarian Syndrome, Hypothyroidism, Viral Hepatitis, Chronic obstructive pulmonary disease

T2DM subgroup adjusted factors: Hypertension, Abnormal lipid metabolism, Hypertension diagnoses, Using hypertension treatment, Hyperlipidemia, Using adjust lipid agents, Hypertension and abnormal lipid metabolism, Abnormal liver function, Cirrhosis, Chronic kidney disease, Obstructive Sleep Apnea, Polycystic Ovarian Syndrome, Hypothyroidism, Viral Hepatitis, Hp. Infection, Chronic obstructive pulmonary disease

| Risk factors | Extrahepatic Cancers Yes | Extrahepatic Cancers No | OR (95% CI) | *P* value |
|---|---|---|---|---|
| **Age<30** | 105 | 5,979 | **Reference** | |
| **Age 30 to 39** | 380 | 16,111 | **1.53 (1.23,1.90)** | ***<0.001*** |
| **Age 40 to 49** | 913 | 20,855 | **2.89 (2.35,3.55)** | ***<0.001*** |
| **Age 50 to 59** | 2,147 | 27,536 | **4.86 (3.97,5.93)** | ***<0.001*** |
| **Age 60 to 69** | 1,839 | 16,619 | **6.85 (5.60,8.40)** | ***<0.001*** |
| **Age >70** | 1,221 | 9,947 | **7.95 (6.46,9.78)** | ***<0.001*** |
| **Female** | 3,243 | 32,988 | **1.37 (1.30,1.45)** | ***<0.001*** |
| **Hypertension** | 3,034 | 38,220 | **1.14 (1.08,1.21)** | ***<0.001*** |
| **Hyperlipidemia** | 1,666 | 40,069 | **0.56 (0.52,0.59)** | ***<0.001*** |
| **T2DM** | 2,691 | 36,312 | **1.15 (1.08,1.22)** | ***<0.001*** |
| **Drug for hypertension** | 1,183 | 23,036 | **0.71 (0.66,0.77)** | ***<0.001*** |
| **Drug for hyperlipidemia** | 738 | 23,679 | **0.41 (0.38,0.45)** | ***<0.001*** |
| **Drug for T2DM** | 903 | 19,231 | **0.63 (0.58,0.69)** | ***<0.001*** |
| **AST>40U/L** | 1,097 | 15,199 | **1.13 (1.03,1.24)** | ***0.0069*** |
| **ALT>40U/L** | 1,608 | 26,301 | **1.08 (1.00,1.17)** | ***0.0536*** |
| **Cirrhosis** | 50 | 2,011 | **0.38 (0.29,0.51)** | ***<0.001*** |
| **Viral hepatitis** | 233 | 9,526 | **0.37 (0.32,0.42)** | ***<0.001*** |
| **Autoimmune hepatitis** | 7 | 297 | **0.26 (0.12,0.55)** | ***<0.001*** |

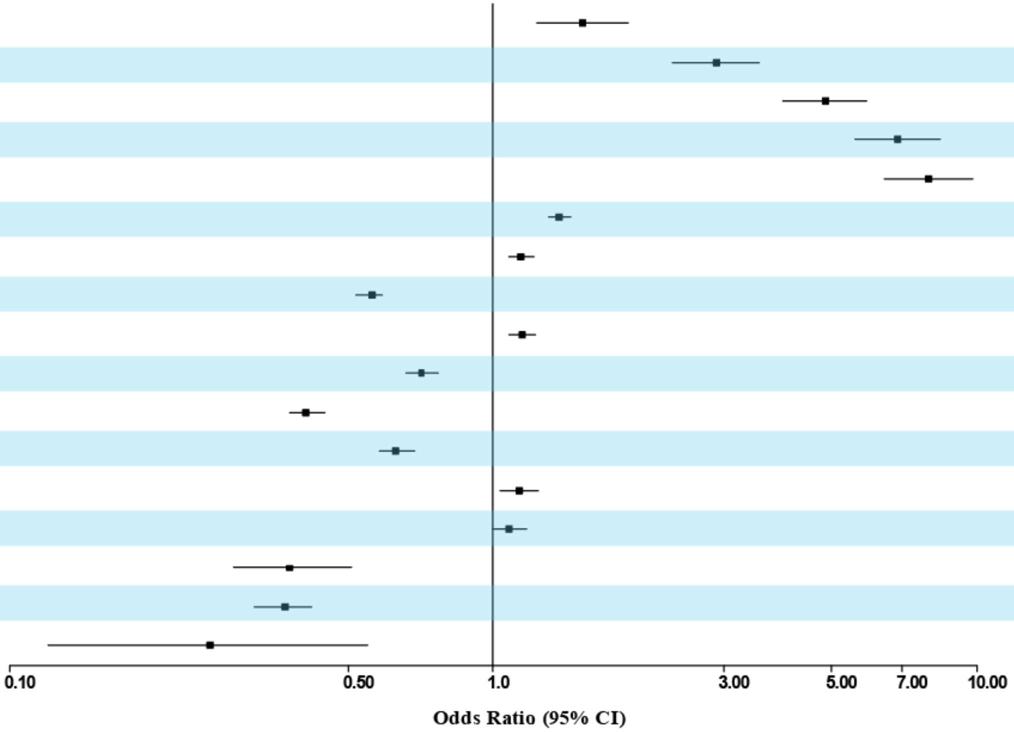

**Supplementary Figure 1**. The association of diagnosed specific metabolic dysfunction, elevated liver enzyme, pharmacological treatments of the metabolic dysfunction, comorbidities and extrahepatic cancers in the MASLD population. ALT: Alanine Aminotransferase; AST: Aspartate Aminotransferase;T2DM: type 2 diabetes mellitus

| Risk factors | Extrahepatic Cancers Yes | Extrahepatic Cancers No | OR (95% CI) | *P* value |
|---|---|---|---|---|
| **Age<30** | 105 | 5,979 | **Reference** | |
| **Age 30 to 39** | 380 | 16,111 | **1.50 (1.20,1.86)** | ***<0.001*** |
| **Age 40 to 49** | 913 | 20,855 | **2.82 (2.29,3.46)** | ***<0.001*** |
| **Age 50 to 59** | 2,147 | 27,536 | **4.77 (3.91,5.83)** | ***<0.001*** |
| **Age 60 to 69** | 1,839 | 16,619 | **6.82 (5.57,8.35)** | ***<0.001*** |
| **Age >70** | 1,221 | 9,947 | **8.10 (6.58,9.96)** | ***<0.001*** |
| **Female** | 3,243 | 32,988 | **1.38 (1.31,1.46)** | ***<0.001*** |
| **Hypertension** | 3,034 | 38,220 | **1.11 (1.05,1.18)** | ***<0.001*** |
| **TC over 5.2mmol/L** | 1,420 | 25,029 | **1.01 (0.92,1.10)** | ***0.8938*** |
| **TG over 1.7mmol/L** | 2,401 | 44,356 | **0.83 (0.78,0.89)** | ***<0.001*** |
| **Lower HDL-C** | 2,568 | 39,317 | **0.95 (0.90,1.01)** | ***0.0900*** |
| **LDL-C over 3.4mmol/L** | 1,143 | 21,308 | **0.80 (0.73,0.88)** | ***<0.001*** |
| **T2DM** | 2,691 | 36,312 | **1.13 (1.06,1.20)** | ***<0.001*** |
| **Drug for hypertension** | 1,183 | 23,036 | **0.72 (0.67,0.78)** | ***<0.001*** |
| **Drug for hyperlipidemia** | 7,38 | 23,679 | **0.36 (0.33,0.39)** | ***<0.001*** |
| **Drug for T2DM** | 903 | 19,231 | **0.61 (0.56,0.66)** | ***<0.001*** |
| **AST>40U/L** | 1,097 | 15,199 | **1.17 (1.07,1.28)** | ***<0.001*** |
| **ALT>40U/L** | 1,608 | 26,301 | **1.16 (1.07,1.26)** | ***<0.001*** |
| **Cirrhosis** | 50 | 2,011 | **0.41 (0.31,0.55)** | ***<0.001*** |
| **Viral hepatitis** | 233 | 9,526 | **0.40 (0.35,0.46)** | ***<0.001*** |
| **Autoimmune hepatitis** | 7 | 297 | **0.28 (0.13,0.59)** | ***<0.001*** |

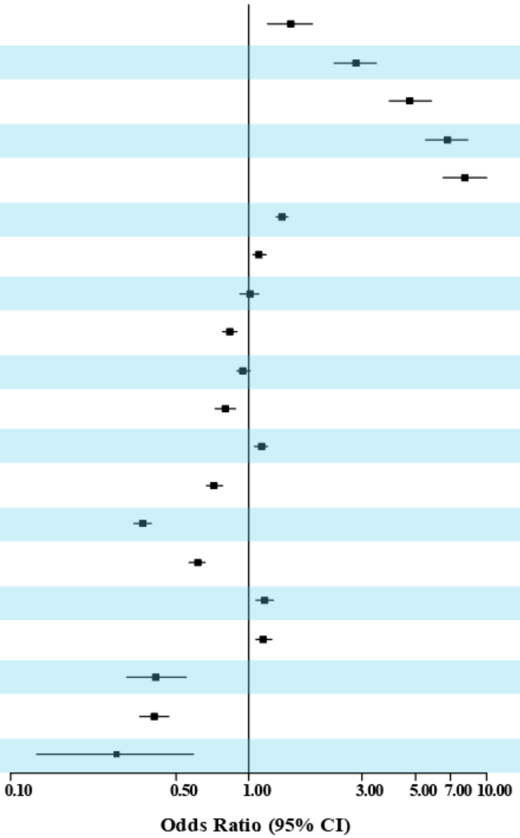

**Supplementary Figure 2**. The association of diagnosed specific dyslipidemia, elevated liver enzyme, pharmacological treatments of the metabolic dysfunction, comorbidities and extrahepatic cancers in the MASLD population. ALT: Alanine Aminotransferase; AST: Aspartate Aminotransferase; T2DM: type 2 diabetes mellitus; TC: Total cholesterol; TG: Triglyceride.

| Risk factors | Extrahepatic Cancers | | OR (95% CI) | *P* value |
|---|---|---|---|---|
| | Yes | No | | |
| **Age<30** | 105 | 5,979 | **Reference** | |
| **Age 30 to 39** | 380 | 16,111 | **1.39 (1.12-1.74)** | ***0.0030*** |
| **Age 40 to 49** | 913 | 20,855 | **2.54 (2.06-3.12)** | ***<0.001*** |
| **Age 50 to 59** | 2,147 | 27,536 | **4.23 (3.46-5.17)** | ***<0.001*** |
| **Age 60 to 69** | 1,839 | 16,619 | **5.79 (4.72-7.10)** | ***<0.001*** |
| **Age >70** | 1,221 | 9,947 | **6.38 (5.18-7.86)** | ***<0.001*** |
| **Female** | 3,243 | 32,988 | **1.40 (1.33-1.48)** | ***<0.001*** |
| **Abnormal liver function** | 509 | 10,340 | **0.88 (0.80-0.97)** | ***0.0073*** |
| **Cirrhosis** | 50 | 2,011 | **0.49 (0.37-0.66)** | ***<0.001*** |
| **CKD** | 258 | 7,388 | **0.46 (0.41-0.53)** | ***<0.001*** |
| **CVD** | 3,445 | 44,816 | **0.86 (0.81-0.91)** | ***<0.001*** |
| **OSA** | 41 | 1,516 | **0.45 (0.33-0.62)** | ***<0.001*** |
| **PCOS** | 6 | 319 | **0.65 (0.29-1.48)** | ***0.3101*** |
| **Viral hepatitis** | 233 | 9,526 | **0.44 (0.38-0.50)** | ***<0.001*** |
| **Hp infection** | 83 | 1,805 | **0.66 (0.53-0.82)** | ***<0.001*** |
| **Autoimmune hepatitis** | 7 | 297 | **0.32 (0.15-0.68)** | ***0.0029*** |
| **COPD** | 134 | 874 | **1.58 (1.31-1.90)** | ***<0.001*** |

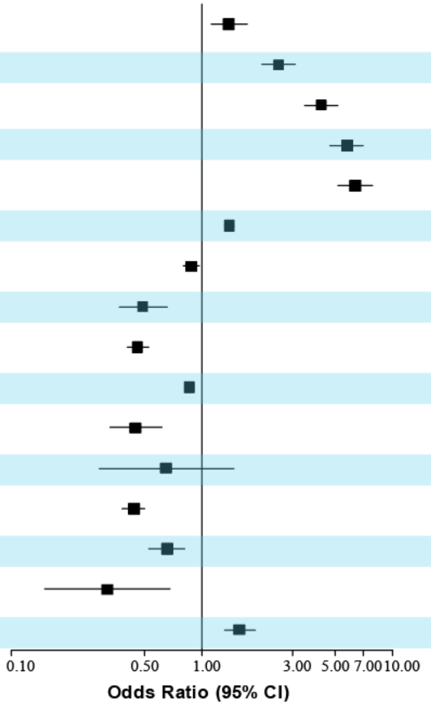

**Supplementary Figure 3**. The association of comorbidities and extrahepatic cancers in the MASLD population. CKD, Chronic kidney disease, OSA, Obstructive Sleep Apnea, PCOS, Polycystic Ovarian Syndrome, COPD, Chronic obstructive pulmonary disease.
